# A Preliminary Characterization of Canonicalized and Non-Canonicalized Section Headers Across Variable Clinical Note Types

**DOI:** 10.1101/2021.03.13.21253523

**Authors:** Junjie Wang, Shun Yu, Anahita Davoudi, Danielle L. Mowery

## Abstract

In the electronic health record, the majority of clinically relevant information is stored within clinical notes. Most clinical notes follow a set organizational structure composed of canonicalized section headers that facilitate clinical review and information gathering. Standardized section header terminologies such as the SecTag terminology permit the identification and standardization of headers to a canonicalized form. Although the SecTag terminology has been evaluated extensively for history & physical notes, the coverage of canonical section header terms has not been assessed across other note types. For this pilot study, we conducted a coverage study and characterization of canonical section headers across 5 common, clinical note types and a generalizability study of canonical section headers detected within two types of clinical notes from Penn Medicine.

## Introduction

Unstructured notes are a rich source of clinically important information, some not otherwise found among the structured data fields of the electronic health record (EHR). The types of clinical notes generated at the point of care can vary by specialty (radiology vs. emergency department (ED) reports), writer (physician vs. nurse), timeline of events (discharge summary vs. progress note), length (short telephone encounter vs. long history & physical (H&P) notes), among other comparisons. These clinical notes can be documented using a predefined template, which organizes relevant clinical information into subsections designated by section headers. In some cases, clinicians can create new sections to organize content into novel sections. These templates, initially designed to be used by clinicians to provide a cognitive framework for their clinical assessments, can be leveraged to assist in information gathering.

In a recent literature review, Pomares-Quimbaya et al. defines a section as “*a text segment that groups together consecutive clauses, phrases, or sentences that share the description of one dimension of a patient, patient’s interaction, or clinical findings* [1].” The sections used vary for the same reasons that clinical note types vary, e.g., variable concept specificity, clinical content, narrative structure, and term synonymy. In particular, clinical notes (e.g., progress notes) can follow the traditional, higher-level SOAP (e.g., “subjective”, “objective”, “assessment”, “plan”) labels [2,3] or more precise, lower-level labels (e.g., “chief complaint” is a type of “subjective” section). Some section headers (e.g., family history) are commonly observed across notes types (e.g., H&P and ED notes); other sections (e.g., specimens) are more frequently associated with a particular note type (e.g., laboratory report). Some sections are explicitly denoted with a case, punctuation, line breaks, or white spacing [4,5]; other sections are implied by either topical [6], temporal [7], or locational shifts in the narrative. In practice, a canonical set of section headers are not often standardized across institutions, or sometimes even within a department, leading to multiple section variants with synonymous meaning (e.g., “family history”, “FHx”). Although there’s no formalized agreement of what constitutes a section segment, there is consensus that identifying the section segmentation for an extracted clinical event can have implications for determining whether that event occurred (actual vs. hypothetical), who experienced the event (family vs. patient) and when it occurred (distant past, recent past, current encounter, future) among other assertions [1, 8].

Although section header terminologies i.e., SecTag, have been developed and evaluated for their coverage of section headers for more general, summarizing notes, e.g., discharge summaries [9] and H&P notes [4], it has not been assessed for coverage of section headers for other semantically-rich, high-value note types e.g., writer-specific note types (e.g., physician and nursing notes) as well as domain-specific note types (e.g., radiology and echocardiograms). Additionally, there has been no characterization of non-canonicalized section headers that could provide semantically-meaningful information for extracting clinical events e.g., problem-oriented section headers (“lung cancer:”). These non-canonicalized section headers could be valuable for improving information retrieval, extraction, and summarization efforts for clinical and translational research efforts e.g., developing natural language processing (NLP)-powered applications for identifying relevant patient cohorts for genotype/phenotype association studies, clinical trial recruitment, and clinical decision support among other use cases.

Our long-term goal is to develop an NLP-powered, section tagger to identify and canonicalize explicit, semantically-meaningful, and high-value section headers across a variety of note types for patient cohort identification. Our most immediate use case is a translational research study for a lung cancer cohort. Our short-term goal, in this pilot study, is to 1) conduct a coverage study of canonical section headers using a standardized, section header terminology, 2) characterize the themes of section headers that did not canonicalize, and 3) assess the generalizability of the section terminology to University of Pennsylvania notes to inform future, automatic section header identification efforts for an ongoing translational research study.

## Methods

In this Institutional Review Board-approved study, we queried notes from the Multiparameter Intelligent Monitoring in Intensive Care (MIMIC)-III database [10]. The MIMIC-III database contains a diverse and comprehensive collection of clinical data for patients admitted to the Beth Israel Deaconess Medical Center in Boston, Massachusetts. MIMIC-III is a relational database consisting of 26 tables with information related to admissions including patients, caregivers, diagnoses, procedures, laboratory events, and other coded data as well as the associated clinical notes. We focused our study on clinical note types that typically contain explicit, but variable section headers commonly documented across institutions. For this study, we processed only - discharge summary, echocardiogram, nursing, physician, and radiology notes - codified and stored in the MIMIC-III noteevents table.

### SecTag Terminology

One of the most comprehensive section header terminologies was developed at Vanderbilt University using H&P notes. This section terminology is leveraged by SecTag, a section tagger that integrates spelling correction, hierarchical terminology rules, and a naive Bayesian scoring method to detect sections from clinical notes [11,12]. To date, the SecTag terminology contains 1129 canonical section labels (e.g., chief_complaint), 6773 section variants (e.g., “cc”, “chief complaint”), and an average of 6 synonym variants per canonical section label^1^.

### Coverage of Canonicalized and Characterization of Non-canonicalized Section Headers

For 10,000 randomly-sampled notes (2,000 of each note type), we report the relationship between the average number of canonicalized, candidate headers and tokens by note type. We depict the distribution of all matched (canonicalized) and unmatched (non-canonicalized) section headers compared against the SecTag terminology across notes types. We further report the distribution of unique matched and unmatched section headers as well as the top 5 most common, unique section headers (unique lexical header variants observed once special characters were removed and case was converted to sentence case) across all notes.

### Review and Annotation of Candidate Section Headers

For notes with unmatched headers, we randomly-sampled and reviewed 50 notes (10 clinical notes x 5 note categories) to determine whether the candidate header should have been encoded using following steps (**Figure 1**):

**Figure 1.**
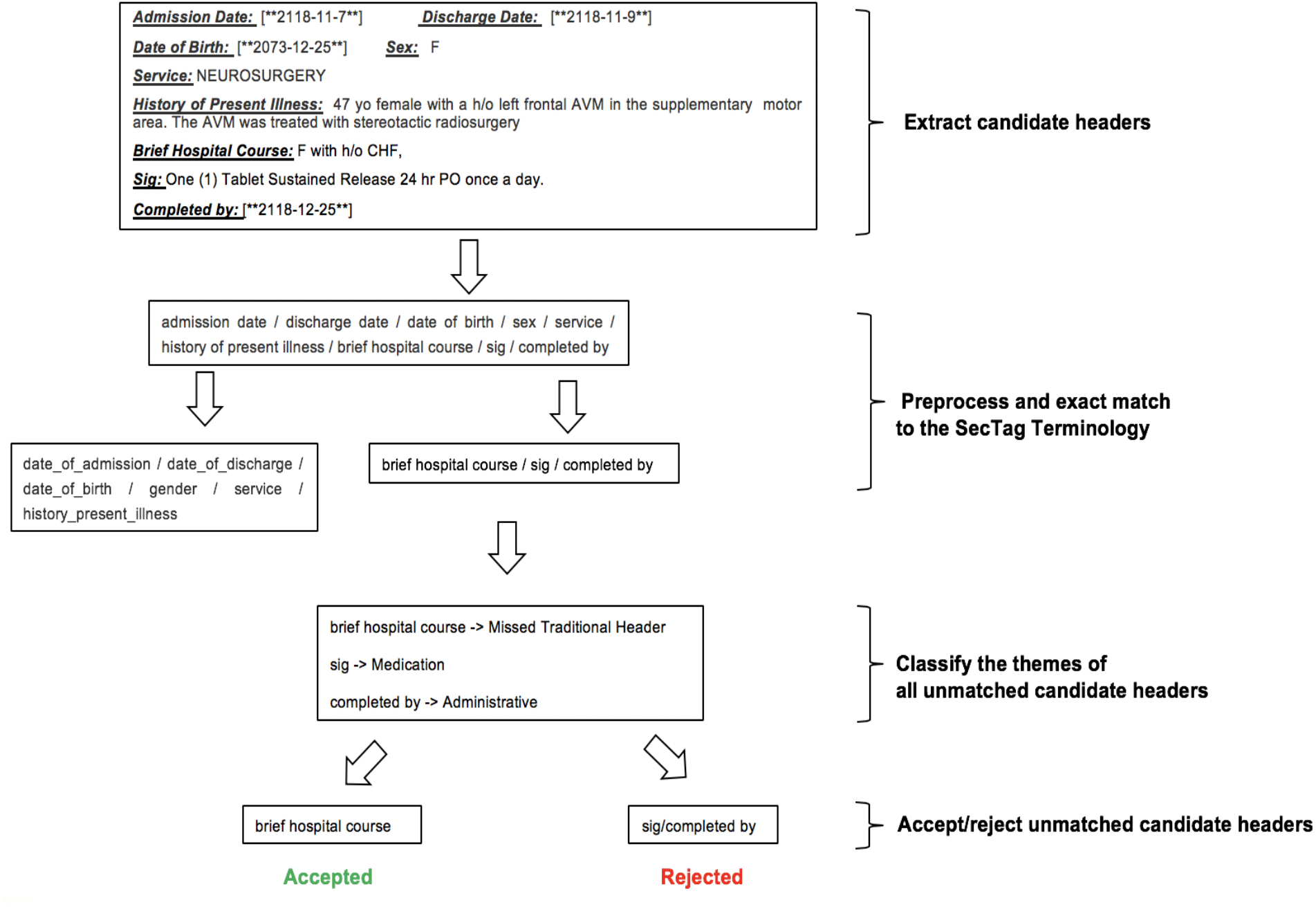
Flowchart of review and characterization of candidate section headers

➢ **<u>Step 1</u>**: Generate a list of candidate section headers using regular expressions to identify phrases likely signifying a section. We created sensitive regular expressions to identify sections denoted as 1 or more words followed by a colon (:) or hash (-) then a space (“Family History: “). These regular expressions do not rely on line breaks because notes from our data warehouse are not stored with these section boundary delineators.
➢ **<u>Step 2</u>**: Match all candidate headers to the SecTag terminology by removing special characters from both the candidate header and canonical header before applying an exact match criterion i.e., same tokens in each set.
➢ **<u>Step 3</u>**: Generate a list of all unmatched candidate headers and their associated contents. Typically, we determine the window size to be 100 characters unless the text that precedes or follows the header is shorter.
➢ **<u>Step 4</u>**: Classify the themes of all unmatched candidate headers as one of 9 types:
  - *missed traditional header*, phrase is missing from the terminology, e.g., “Other medications:”
  - *procedure*, device, treatment, or method used to treat a patient, e.g, “arterial line:”
  - *question/answer*, Q/A response syntax, e.g., “Any signs of infection at the wound site:”
  - *administrative*, clerical information, e.g., “Dictated By:”
  - *additional assertions*, presence/absence of events, e.g., “All the following are negated:”
  - *problem-specific*, disease, disorders, observations, e.g., “Hypertension:”
  - *medication*, related to drugs, their modifiers or administration/usage, e.g., “Refills:”
  - *need previous header to contextualize*, phrase required parent header to interpret, e.g., “Total In:”
  - *hyphenated word*, e.g., compounded words spuriously identified, e.g., “right-sided rib fracture”
➢ **<u>Step 5</u>**: Accept or reject each unmatched and classified candidate header. We <u>accepted</u> high-value sections that easily generalize across institutions and provide semantically important information necessary for accurately representing, encoding, and extracting clinical events including signs, symptoms, findings, observations, risk factors, diagnoses, etc. For now, we <u>rejected</u> candidate headers that we believed would be low-value, not likely to contain clinically meaningful events and are not administrative or encoded as structured fields in the EHR.

Finally, we present the distribution of accepted and rejected unmatched candidate headers and themes described across the note types.

### Demonstration of Generalizability

To assess the generalizability of the SecTag terminology on Penn Medicine notes, we randomly-sampled 50 notes of 2 types (discharge summary and radiology notes) from an advanced lung cancer cohort study for which we aim to target specific sections for extracting lung nodule progression status from radiology reports as well as pertinent past medical diagnoses, diagnostic and therapeutic histories, family histories, and social histories from discharge summaries. SY, an oncologist, annotated all high-value headers from these notes. AD applied the section terminology using the NLP pipeline to each note. Compared to the reference standard headers annotated by SY, AD determined correctly identified section headers (true positives), incorrectly omitted high-value section headers (false negatives), and spuriously matched candidate headers (false positives). We report the average recall, precision, and F1-score of the candidate header algorithm using exact and partial string matching and the errors commonly generated from this dataset.

## Results

Of the 10,000 notes from the sample set (2,000 of each note type), the average number of canonicalized, candidate headers (h) and tokens (t) by note types were noted as: discharge summaries (h: 60, t: 1564), nursing notes (h: 14, t: 218), physician notes (h: 74, t: 730), radiology notes (h: 7, t: 202) and echocardiograms (h: 23, t: 332). (**Figure 2**).

**Figure 2.**
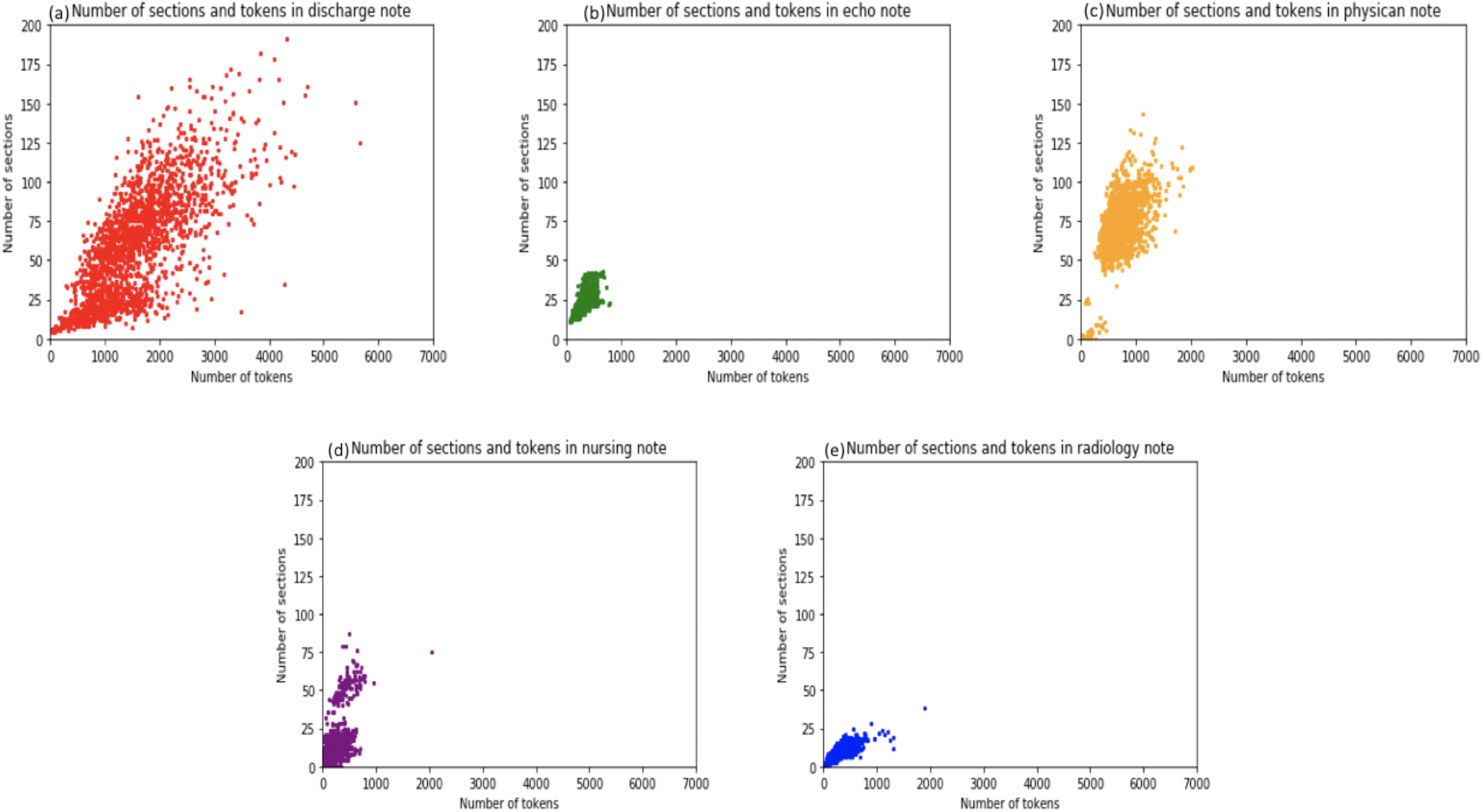
Number of sections and tokens: **a**. discharge summary, **b**. nursing, **c**. physician, **d**. radiology, **e**. echo.

### Coverage Study of Canonicalized Section Headers

First, we attempted to match all candidate section headers within the sample set to the SecTag terminology. The proportion of candidate section headers that matched to a canonical header within the SecTag terminology varied across note types. For discharge summaries, 53.3% of candidate header mentions match to SecTag terminology; in contrast, only 26.4% of echo candidate header mentions match to SecTag terminology (**Figure 3a**). A review of all unmatched, unique candidate header variants revealed the following distribution by note types: radiology (93%), echocardiogram (92.1%), discharge summary (89.8%), physician (89.1%), and nursing (80.3%) notes (**Figure 3b**).

**Figure 3.**
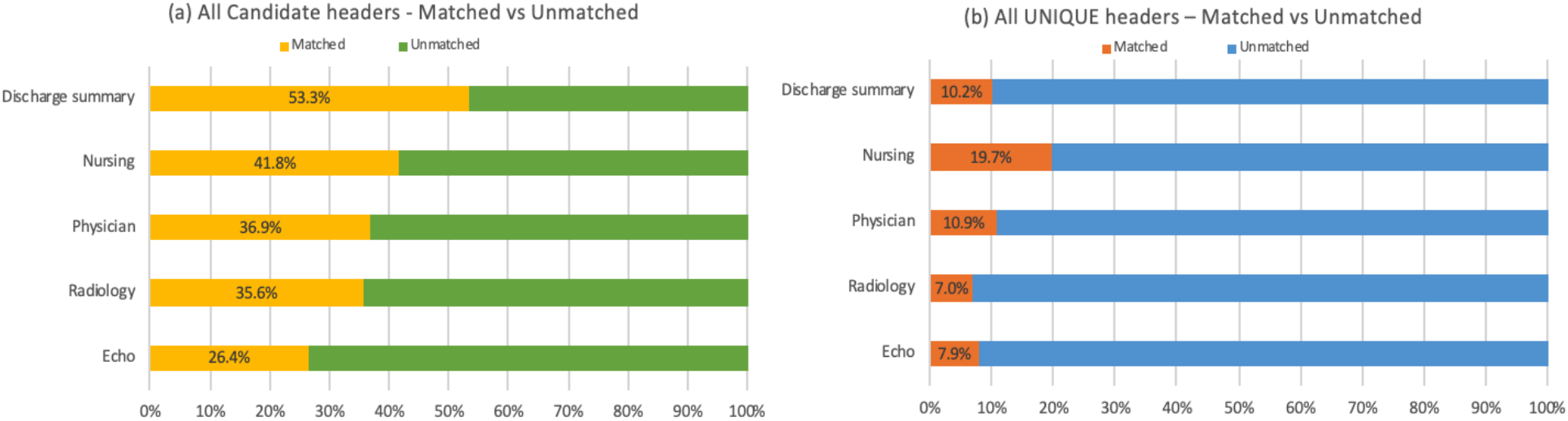
Matched and unmatched candidate headers: (a) all instances and (b) unique headers across note types

### Characterization of Non-canonicalized Section Headers

Of all unique header variants, we enumerate the top 5 most common and depict the distribution of unmatched (**Figure 4a-e**) and matched (**Figure 4f-j**) candidate headers. The unmatched headers were classified by themes, then either accepted or rejected based on their value for supporting clinical and translational research.

**Figure 4.**
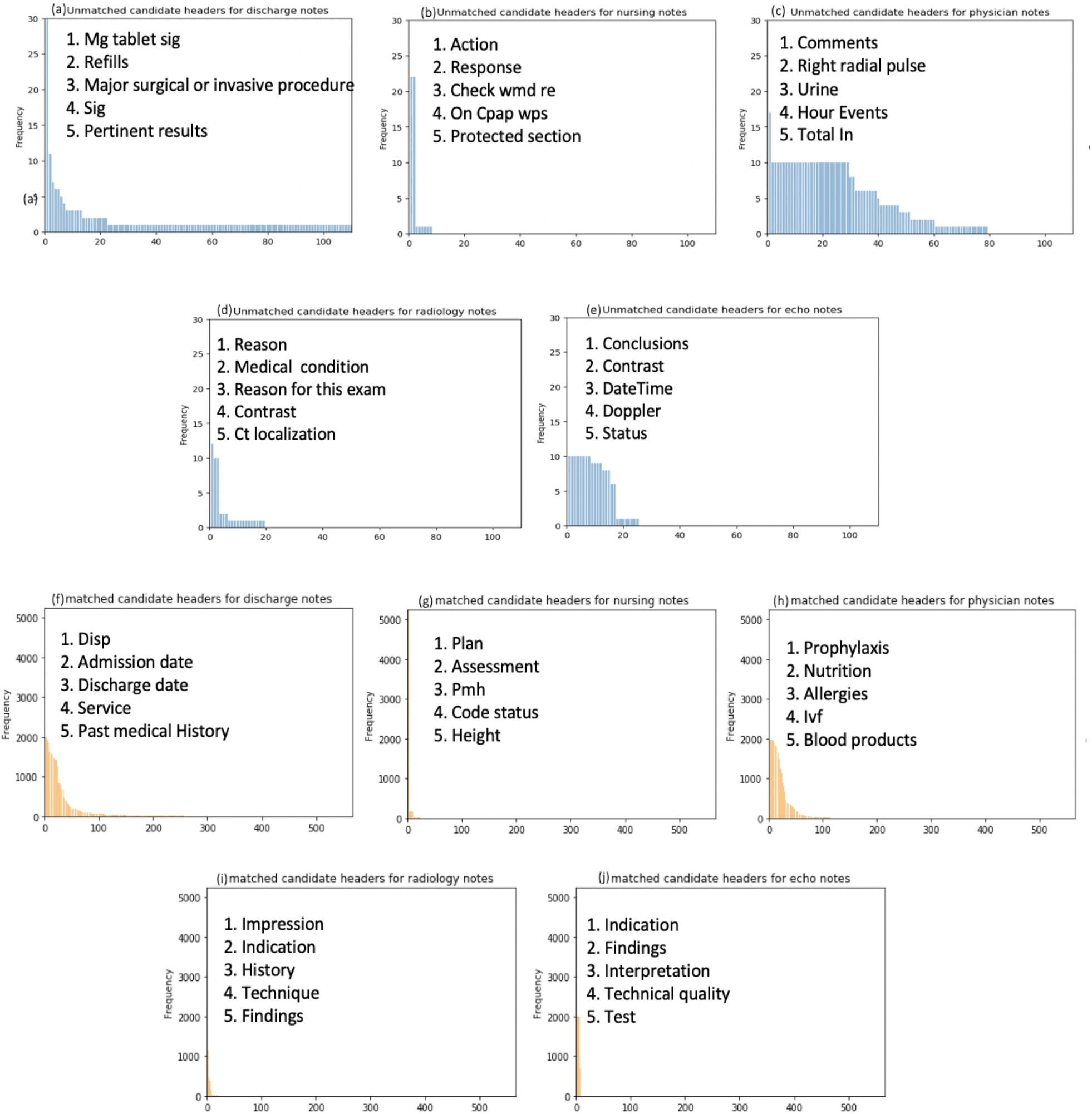
Top 5 unique, **unmatched** headers: **a**. discharge summary, **b**. nursing, **c**. physician, **d**. radiology, **e**. echo. Top 5 unique, **matched** headers: **f**. discharge summary, **g**. nursing, **h**. physician, **i**. radiology, **j**. echo. Unmatched header short forms: sig=signature, cpap=continuous positive airway pressure, wmd=weighted mean difference, ct=computed tomography. Matched header short forms: disp=disposition, pmh=past medical history, ivf=intravenous fluid.

In **Figure 5**, for our review of 50 notes, we show a sunburst infographic to depict the relationships between the unmatched unique header variants, characterization theme, note type, and their acceptance or rejection (outer-to-innermost layer). We’ll review this infographic from the inner-to-outermost layer. In our sample dataset, containing 50 clinical notes, we accepted 64% of the unmatched candidate headers and rejected 46% of them. For each note type, the proportion of accepted (a) and rejected (r) candidate sections were as follows: discharge summary (a: 36.3%, r: 63.7%), radiology (a: 90.1%, r: 9.9%), echo (a: 86.9%, r: 13.1%), nursing (a: 88.0%, r: 12.0%), and physician (a: 66.3%, r: 33.7%) notes. The most commonly accepted themes by note types were: discharge summary (procedure, problem-specific, traditional), radiology (traditional and procedure), echo (anatomical location, traditional), nursing (traditional), and physician (procedure, problem-specific, traditional) notes. The most commonly rejected themes by note types were: discharge summary (medication, administrative, procedure), radiology (procedure), echo (procedure, administrative), nursing (administrative), and physician (procedure, administrative, medication) notes.

**Figure 5.**
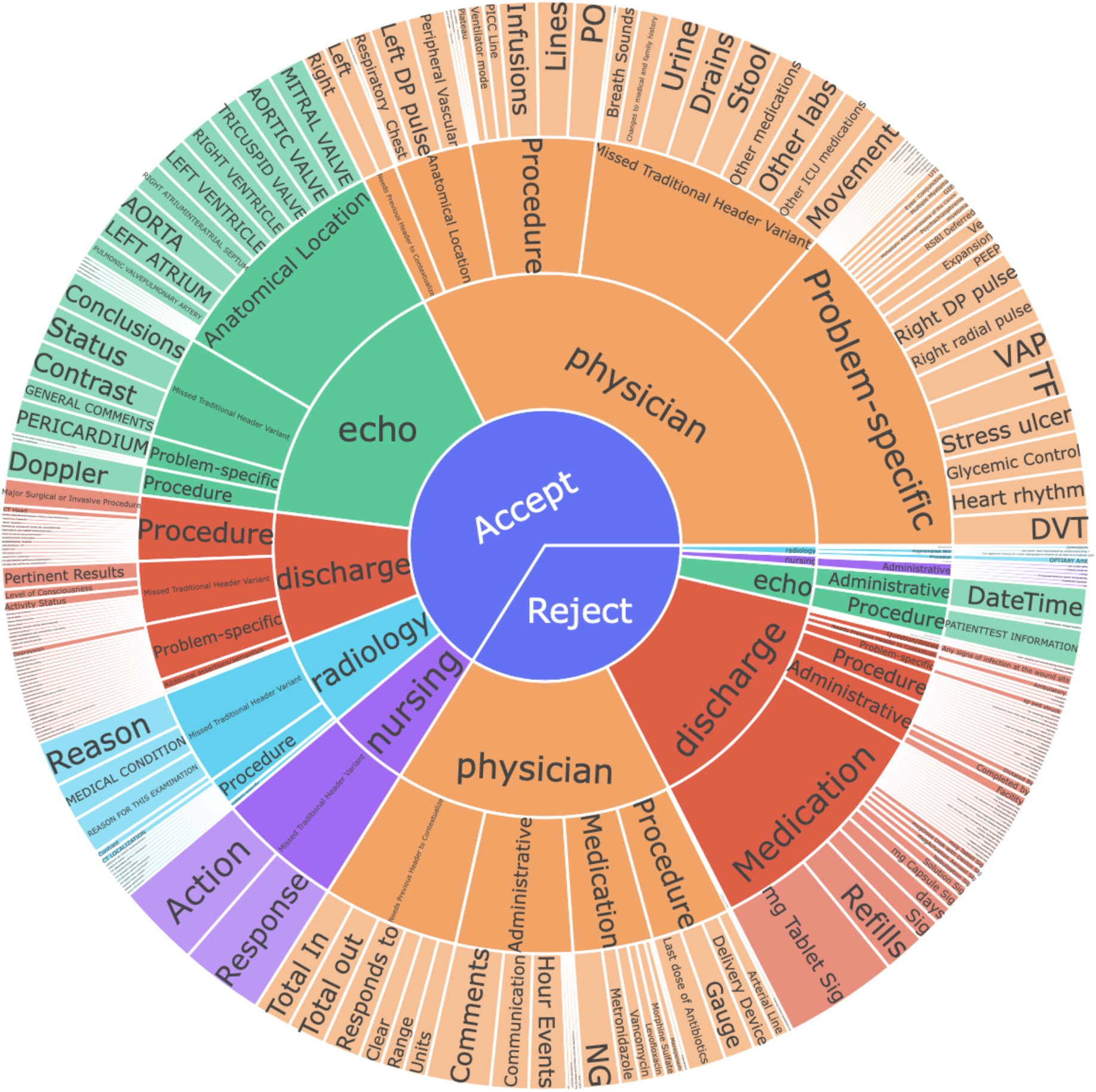
Sunburst of unmatched candidate sections (inner-to-outermost layer): accepted/rejected > note type > theme type > all candidate headers

### Demonstration of Generalizability

In **Table 1**, using the NLP pipeline, we applied the SecTag terminology to Penn Medicine discharge summaries and radiology notes which are pertinent to an ongoing lung cancer cohort study. The NLP pipeline identified about twice as many (more or less) section headers than annotated within the reference standard for both note types. For discharge summaries, by relaxing the match criteria from exact to partial, we observed an improvement in average recall (0.30 to 0.88) and precision (0.15 to 0.62). For radiology notes, although relaxing the match criteria did not result in a change in average recall (0.80), we did observe improvements in average precision (0.56 to 0.73).

**Table 1.** Performance of section tagger on sample (50 notes x 2 types = 100 notes) from a lung cancer cohort.

| Note type | Average exact match |  |  | Average partial match |  |  | Annotation counts |  |
| --- | --- | --- | --- | --- | --- | --- | --- | --- |
|  | Recall | Precision | F1-score | Recall | Precision | F1-score | Reference Standard | NLP section pipeline |
| *Discharge Summaries* | 0.30 | 0.15 | 0.20 | 0.88 | 0.62 | 0.71 | 183 | 323 |
| *Radiology Notes* | 0.80 | 0.56 | 0.65 | 0.80 | 0.73 | 0.84 | 54 | 117 |

## Discussion

Our goal was to conduct a coverage study of canonicalized section headers using a standardized, section header terminology and characterize the themes of non-canonicalized section headers to inform future, automatic section header identification efforts. Using lessons learned from this preliminary study, we will develop an automated section tagger to identify and canonicalize each high-value, explicit section across multiple note types. After canonicalizing these clinical headers, clinical notes can be stored structured in a clinical database and queried by their canonical headers. This will support a more precise query of salient clinical events extracted by NLP for identifying relevant patient cohorts for clinical research studies. This study of canonicalized and non-canonicalized section headers from five clinical note types aims to improve our understanding of additional note type-specific, semantically-important, and high-value section headers that are not commonly being addressed across existing section tagging efforts in the literature. Of the 10,000 notes we sampled and identified an exact match of section variants from the SecTag terminology, we observed a positive association between the number of sections and tokens of discharge summaries, i.e., the more content in a note type, the more sections and tokens. Physician notes and discharge summaries had the highest number of sections and token counts, which was not surprising given the amount of clinical information documented relevant to the course of the patients’ visit.

### Coverage Study of Canonicalized Section Headers

First, we applied an exact match criterion using the SecTag terminology. We observed a moderate proportion of section headers that were not canonicalized to the SecTag terminology ranging from 46.7% to 73.6%. Once each section header variant was standardized to identify unique headers (same case, no special characters), the proportional distribution of unmatched headers rose ranging from 80.3% to 93.0%. The relative proportions of matched-to-unmatched between all candidate headers and unique headers have a general trend across note types except for discharge summaries. In discharge summaries, 10.2% of unique headers account for 53.3% of all candidate headers. This finding suggests that there are a higher variability or more synonyms for section headers used within discharge summaries over other note types. Surprisingly, nursing notes do not follow the same trend. We hypothesize that for other note types like radiology and echocardiograms, we would anticipate some degree of template usage or fewer synonyms for section headers resulting in lower variability.

### Characterization of Unmatched Section Headers

In total, there are 899 unmatched headers. For those notes with unmatched headers, we randomly sampled 20 notes from 5 note types (n=100 notes) to characterize common themes amongst unmatched headers. For all note types, we observed traditional section headers or synonyms that were missing from the list of canonicalized sections in the SecTag terminology (e.g., “level of consciousness” should exist for SecTag kmname level_of_consciousness). For several note types, section headers denote semantically-important and high-value information (problems, procedures, and anatomical locations) that could be important for extracting clinical events. For discharge summaries, physician, and radiology notes, synonyms for clinical problems were not found. This makes sense given discharge summaries and physician notes can be written using the problem-oriented medical record format (POMR) in which each clinical problem (e.g., “pneumonia:”) is enumerated with associated clinical details. This is often done in the intensive care unit (ICU) setting. Radiology notes contain missing headers for components related to procedures and investigational methods necessary to identify critical findings (e.g., “contrast:”) and echocardiograms contain missing anatomical location headers (e.g., “aorta:”) that convey where a pertinent clinical finding was observed during the review of cardiovascular structures. These observations signify opportunities for novel expansion upon the existing literature for supporting the canonicalization of section headers across various clinical note types.

### Demonstration of Generalizability

We applied and evaluated the SecTag terminology on Penn Medicine discharge summaries and radiology notes using the NLP pipeline. Using an exact match criterion, the NLP pipeline did not detect many headers due to a missing colon at the end of the phrase. In particular, a considerable number of false negatives contributed to initial low recall for discharge summaries. This finding is not surprising given our initial characterization revealed that discharge summaries have notably more sections than most other note types, and therefore, more opportunities for omissions. Some omitted sections include domain-specific history headers (“Onc history”), problem headers (“HTN”, “Thrush”), and subsections. To address domain-specific history headers, we could include a Jaccard similarity comparison to existing high-value and commonly documented headers. To encode problem-oriented headers, we could leverage biomedical vocabularies i.e., the Unified Medical Language System (UMLS). To readily identify subsections in the future, we could include knowledge of the prior recognized section header and encode textual features to capture and signal an ontological relationship with the next semantically meaningful noun phrase following the parent header. Conversely, false positives were also generated based on the colon in the regular expression for capturing candidate headers, resulting in low precision. Low-value sections spuriously tagged include medication-related contexts (“hours total dose”) and administrative headers (“attending signature”). We hypothesize that these section headers could be omitted through simple post-processing using high precision term lexicons. Using a partial match criterion, we observed an increase in all the metrics (recall, precision, F1-score), most notably for discharge summaries. By relaxing the matching span (e.g., detecting “allergies” rather than the full section header “allergies drug”), we improved coverage. Practically-speaking, even partial matches may still capture enough semantics to support a given information extraction task (e.g., extracting sources of allergic reactions). Additional approaches to improve candidate header identification include adjusting steps for pre-processing the notes or post-processing the candidate headers i.e., cleaning the texts, removing empty spaces, etc. For this initial assessment, we applied a top-down approach, applying an existing section terminology. In addition to these enumerated approaches, we will experiment with bottom-up approaches that leverage deep learning to incorporate document structure as well as prior knowledge of the header sequence, note type, and writer to improve section header detection.

## Limitations

Although we have discovered new knowledge for developing strategies for automatically identifying section taggers for a variety of clinical notes, our study has several limitations. Given the scope of our project, we rejected medications (for now) and low-value, administrative section headers. Additionally, we also omitted some seemingly valuable header themes (e.g., “total in:”, “total out:”) which don’t provide inherent value without clinical context (e.g., “fluids:”). These information types are also fairly consistently stored as structured fields. This decision was driven by the lack of immediate need to support our clinical research studies. We recognize that these determinations can be use case driven and can entail some subjectivity on the part of the reviewer as do most annotation tasks. Although we focused our work on only 5 note types, we selected note types that are commonly processed to support research studies and capture a diverse set of writers, specialties, and intentions. As an initial step, we applied the SecTag terminology to Penn Medicine notes to assess the generalizability of the terminology for notes from another academic medical center and non-ICU population. We have not enhanced nor tuned the pipeline to address the nuances of Penn Medicine provider’s documentation styles. A much larger training and testing set is being developed to experiment with the aforementioned solutions, and create a robust, section tagger to address these initial findings in the months to come.

## Conclusions

The current study provides an observational study to inform the development of an NLP module for identifying and canonicalizing, high-value section headers within clinical notes. The classification of unmatched section headers on sample notes and initial evaluation of the terminology on Penn Medicine notes gives us insights for developing a computational approach to further canonicalize, semantically valuable section headers.

## Data Availability

Data is available with proof of permissions through the Medical Information Mart for Intensive Care version 3 (MIMIC-III) database.

https://mimic.physionet.org/

## Acknowledgements

We extend our gratitude to the developers of the SecTag terminology and of the MIMIC-III for making these valuable resources available to the informatics community and to the AMIA reviewers for the thoughtful feedback on this preliminary work. This project was partially funded by the Institute for Biomedical Informatics/Abramson Cancer Center emerging Cancer Informatics Center of Excellence pilot project.

## Footnotes

1 https://vumc.org/cpm/sites/vumc.org.cpm/files/public_files/sec_tag.zip

